# Pathways to mental health services across local health systems in sub-Saharan Africa: Findings from a Systematic Review

**DOI:** 10.1101/2024.01.11.24301103

**Authors:** Samuel Adeyemi Williams, Mamadu Baldeh, Abdulai Jawo Bah, Dimbintsoa Rakotomalala Robinson, Yetunde C. Adeniyi

## Abstract

Mental illnesses significantly affect patients, families, and their communities. The different pathways to care, both formal and informal, influenced the timing and appropriate care. This literature review identifies pathways to access mental health services and suggests a collaborative model for mental health care across sub-Saharan Africa. We systematically searched multiple databases for studies reporting on primary qualitative and quantitative studies on pathways to mental healthcare services across Sub-Saharan Africa. Descriptive analysis of pathway stages was done according to Goldberg and Huxley’s ’model of Levels and Filters’. Overall, twenty-nine We included 29 studies in the final review. Biomedical services were the preferred treatment option. The majority (70%) used traditional and religious healers as the first point of mental health care. The median duration for the delay in seeking treatment in a health facility was six and fifty-four months. Patients who sought care from traditional and faith healers were found to have experienced the most prolonged delay without treatment. This study emphasizes that the call for collaboration between the two care systems can no longer be ignored. A proposed new model for collaboration between biomedical and traditional /faith healers that focuses on education and adopting a new referral framework.

## Introduction

Mental health (MH) has been a low priority in most low and middle-income countries (LMIC), as most healthcare programs focus on infectious and non-communicable diseases (Prince et al., 2007). Although there has been a growing awareness of the importance of mental health as a critical component of health in general, especially in child development, probably due to the tremendous change in children’s and adolescents’ health and disease patterns (Prince et al., 2007; Patel et al., 2007), MH is still left out of most policy agendas across Sub-Saharan Africa (SSA) due to competing priorities with other healthcare demands, poverty, and conflicts. This is also reflected in the World Health Organization (WHO) Mental Health Atlas 2022, which shows considerable disparities in resource allocation (financial and human) for mental health in LMIC. The estimated average government MH budget is only around 2% of the total health budget (WHO, 2020). Moreover, 60% of this expenditure is directed towards outdated approaches in psychiatric hospitals, with an out-of-pocket expenditure of over 40% (Mental health atlas, 2017).

The global burden of mental health disorders varies mainly due to the heterogeneity in disease classification and methods used in measurement (Vigo et al., 2016). The WHO estimates a worldwide prevalence of 14% of the global burden of diseases attributable to mental disorders, with 75% of people affected in low-income countries going without adequate treatment (WHO, 2008; Altevogt et al., 2010; Patel et al., 2010). Based on years lived with disability (YLDs) and disability-adjusted life-years (DALYs) measurements of disease burden, Vigo et al., 2016 estimate that mental illnesses account for 32.4% of YLDs and 13% of DALYs, making mental illnesses the most burdensome disease in terms of YLDs. Although these estimates show the extent of the global burden of mental illnesses over the years, the much-needed attention from stakeholders to place appropriate interest in prioritising funding and treatment for persons with mental disorders is almost inexistent, especially across SSA (Bloom et al., 2011) and may therefore lead to neglect, stigma, and discrimination (Saxena et al., 2007; Lasalvia et al., 2013).

In low and medium-income countries including Africa, few skilled professionals are available to cater to the needs of mental health conditions (WHO, 2017). This lack of human resources is not limited to a particular country but is widespread throughout SSA. In Kenya, with a population of 50 million, there are only 45 psychiatrists, and only one is a trained child and adolescent MH professional (WHO, 2017; Kamau et al., 2017). Meanwhile, in Tanzania, there are 0.04 psychiatrists per 100,000 people (WHO, 2017), and in Nigeria, there are 0.09 psychiatrists per 100,000 people. In countries with limited capital and human resources for health, many people resort to seeking care from religious/traditional healers in their quest for healing for many reasons (WHO, 2013). The WHO estimates a ratio of 1:500 traditional/religious healers to the population compared to 1:40,000 doctors/population across Africa (WHO, 2013). This disparity in available human resources, especially at the community level, influences health-seeking behaviour and commonly held traditional beliefs.

An essential component of this study is understanding the patient’s pathways to care through their varied help-seeking behaviours. It will help discern the pathways people take toward care, identify barriers and reasons for the delay, and thus inform policy and practice (Adeosun et al., 2013). Pathways to care for mental disorders refer to the descriptions of care points used before care is sought from a MH professional and the factors that influence the decision-making process (Gater et al., 1991). Rickwood et al. 2012 define help-seeking pathway as the active adaptive process persons take to seek assistance in dealing with mental disorders, and this course is not random but guided by psychological and sociocultural factors (Anderson et al., 2010). To better understand the use of this definition and its application, several theoretical models of pathways to care that predict the decision-making process and investigate factors that guide that process have been proposed in several studies. However, these frameworks or models were designed and focused on the implications in countries with developed health frameworks and well-structured health systems.

Given the recent changes in health service delivery with the introduction of integrated primary care service, which includes MH services, especially across SSA, it is important to review existing knowledge on the patterns of patients’ help-seeking behaviour. A systematic literature review will help identify existing care paths across SSA and understand the influence on timely referral, diagnosis, and treatment. Understanding the pathways to care could provide helpful guidance for public MH efforts to promote awareness (Abdulmalik & Sale, 2012), which will help define a contextual collaborative model between formal and informal MH systems across SSA. It is, therefore, imperative to critically assess the roles different stakeholders play, whether formal or informal, at the time of the first consultation to determine if there is any significant contribution to delay in seeking formal healthcare services and assess the time it takes from experiencing the first symptoms or signs and accessing care. Elucidating on the different pathways to care will help in understanding the barriers that hinder access, including recursive pathways, and identify opportunities for enhancements and collaboration with various stakeholders involved in mental health care (MHC) delivery, including at the community level, is of utmost importance in SSA.

This systematic mapping of pathways to MHC service focused on the pathways to care, perceived cause of mental disorders in SSA and duration on seeking care, and was guided by three research questions:

Research question 1: What are the existing pathways to care for individuals with mental health disorders in SSA?

Research question 2: What socio-demographic characteristics correlate with the pathways to care for people with mental health disorders in SSA?

Research question 3: What is the delay in seeking treatment for mental health disorders in SSA?

In this study, we aim to systematically review existing literature on the pathways to support and psychiatric care in patients with mental disorders across SSA. The findings will, therefore, throw light on help-seeking behaviours related to mental health and inform policymakers on strategies that will improve access to mental health services. Furthermore, these findings will inform the design of a model pathway-to-care approach across SSA.

## Methods

### Search strategy

Following the guidelines outlined in the Preferred Reporting Items for Systematic Reviews and Meta-Analyses (PRISMA), we conducted a comprehensive systematic review to identify the existing pathways to mental health care in practical, real-world settings. Our search approach adhered to the Cochrane Collaboration guidelines and was thorough. With the support of a librarian, mental health experts (Y.C.J. and S.A.W) devised the search strategy for all studies assessing mental health care pathways. Two independent reviewers (M.B. and S.A.W.) examined studies, specifically looking for direct pathways to mental healthcare across SSA, whether documented as primary or secondary findings.

The primary search strategy involved an exhaustive search of academic databases using subject headings and keywords with MeSH terms to find relevant studies. We also hand-searched reference lists of all relevant articles and journals to identify any missed literature that could contribute to the research goal. For any new keyword or term identified, an additional search was done in the databases, and relevant papers were identified until no new article was found. We identified studies on ’pathways to care,’ ’mental health,’ and ’sub-Saharan Africa’ using multiple keywords based on work done by other scholars (Daghash et al., 2019; Latina et al., 2020; Mansfield et al., 2020).

We employed several targeted search strategies, including Booleen operators, to ensure the robustness of the results from different databases (Supporting Information: S1. Search Strategies Legend). We searched the following significant databases: Embase, MEDLINE, CINAHL, PsycINFO and Global Index Medicus.

### Inclusion and Exclusion criteria

We included articles reporting primary empirical studies using various study designs, exclusively on mental health conditions based on all age groups, performed in SSA to explore any pathway to mental health care. We included studies on (i) treatment or help-seeking behaviours for mental health problems and (ii) standardised tools or specific methods to assess pathways to care.

We did not place any limits on the date of the study. We excluded articles that were (i) researched from outside Africa and (ii) published in languages other than English or French (Supporting Information: S2. Table - Selection criteria).

### Quality assessment & Data Extraction

To assess the quality of the studies incorporated in this analysis, the Hong et al. (2018) mixed-method appraisal tool was employed. The methodological quality of three types of research was evaluated: qualitative research, quantitative descriptive studies, and mixed methods studies. Two independent reviewers conducted the quality appraisal process. Percentage scores were utilized to categorize the quality of evidence: (i) 50% indicates low-quality evidence, (ii) 51–75% indicates average-quality evidence, and (iii) 76–100% indicates high-quality evidence.

Two research team members independently screened our records using titles and abstracts. We retrieved the full texts of documents that required further review based on our inclusion and exclusion criteria. We reviewed all the included papers and discussed to reach a consensus in cases of disagreements. In cases where the two reviewers could not decide on an included paper, a third reviewer was consulted to reach a consensus. The degree of concordance among screeners’ findings during the review of abstracts and complete articles was assessed by computing Cohen’s kappa statistics. The interpretation of kappa statistics is as follows: values <0.1 signify no agreement, 0.10–0.20 suggest none to slight agreement, 0.21–0.40 denote fair agreement, 0.41–0.60 indicate moderate agreement, 0.61–0.80 signify substantial agreement, and 0.81–1.00 represent almost perfect agreement.

All references were managed using the Endnote® software. References and PDFs were organised in a specialised folder with comments and annotations. Papers that met the inclusion criteria were examined thoroughly for content familiarisation and extract contributions to the question in a review. Each article involved in the review was critically appraised. The appraisal started with trying to answer the six questions (where, how, when, what, who, and why) developed by Wooliams et al., 2011. For each paper, we extracted (i) publication details: title, author, year, institution, and (ii) descriptive details: Study context, location, mental disorder, pathway to care, and duration to seeking care. Data cleaning and extraction were done using a Microsoft Excel spreadsheet, and descriptive analysis was generated. Both reviewers rechecked extracted data to address any disagreements.

## Data Analysis

We categorised and tallied the study settings and participants, sample size and data collection methods and subsequently presented the data as descriptions and proportions. In order to adequately capture context-specific and sensitive aspects of mental health pathways, a narrative synthesis approach was used to integrate evidence from diverse methodological study designs, including quantitative and qualitative studies addressing pathways to mental health.

We used the adjusted version of the Levels and Filters Model by Goldberg & Huxley, 1996 to categorise our findings. The pathway progression from community to specialised mental health services and the occurrences highlight the transition between various MHC levels. The levels correspond to mental health conditions within the community, those who seek assistance from primary care services, patients who get referred to specialised MH services, and patients who ultimately receive specialised care. These levels are complementary, and analysis was done to reflect the SSA context.

## Results

### Included studies

Table 1 summarises all papers meeting the inclusion criteria over time, study setting and participants, sample size and the data collection methods.

**Table 1.** Included Studies

| Paper | Setting | Study population | Sample size | Methods |
| --- | --- | --- | --- | --- |
| Abdulmalik et al. 2012<br>Pathways to psychiatry care for children and adolescents in a tertiary facility in Northern Nigeria | Nigeria | Children and adolescents | 242 | Interview |
| Bakare et al., 2013<br>Pathway to care: first points of contact and sources of referral among children and adolescent patient seen at a neuropsychiatric hospital in SE Nigeria | Nigeria | Children and adolescents | 393 | Interview |
| Kamau 2017<br>Who seeks child and adolescent mental health care in Kenya? A descriptive clinic profile at a tertiary referral facility | Kenya | Children and adolescents | 166 | Modified WHO Encounter form |
| Abiodun et al. 1995<br>Pathways to mental health care in Nigeria | Nigeria | Adolescents and Adults (>16) | 238 | Modified WHO Encounter form |
| Adeosun et al. 2013<br>The Pathways to the First Contact with Mental Health Services among Patients with Schizophrenia in Lagos, Nigeria | Lagos, Nigeria | Adults | 138 | Semi-structured interview |
| Patel et al. 1997<br>The pathways to primary mental health care in high-density suburbs in Harare, Zimbabwe | Harare, Zimbabwe<br>1997 | Adolescents and Adults (15 – 70) | 53 | Pathways to Care Schedule |
| Temmingh et al. 2008<br>Pathways to care and treatment delays in first and multi-episode psychosis | South Africa | Adults | 71 | WHO Encounter form |
| Odinka et al. 2015<br>The socio-demographic characteristic and patterns of help-seeking among patients with schizophrenia in SE Nigeria | SE, Nigeria | Adults | 367 | WHO Encounter form |
| Nonye et al. 2009<br>Health seeking behavior of mentally ill patients in Enugu Nigeria | Enugu, Nigeria | Adolescents and Adults (15 – 75) | 397 | Interview |
| Lasebikan et al. 2012<br>Social network as a determinant of the pathway to care to mental health service utilization among psychotic patients in a Nigerian hospital | Nigeria | Adolescents and Adults (14 – 58) | 652 | Clinician administered interview |
| Kauye et al. 2014<br>Pathways to care for a psychiatric patient in a developing country | Malawi | Adolescents and Adults (13 – 75) | 128 | WHO Encounter form |
| Girma et al. 2011<br>Patterns of treatment-seeking behavior for mental illnesses in Southwest Ethiopia: a | Ethiopia | Adults | 384 | WHO Encounter form |
| hospital based study |  |  |  |  |
| Appiah-Poku et al. 2003<br>Previous help sought by patients presenting to mental health services in Kumasi, Ghana | Ghana | Adults | 302 | Interview |
| Aghukwa e al. 2012<br>Care seeking and beliefs about the cause of mental illness among Nigerian psychiatric patients and their families. | Nigeria | Adults | 219 | Modified WHO<br>Encounter form |
| Bekele et al. 2008<br>Pathways to Psychiatric Care in Ethiopia | Ethiopia | Children and<br>Adults (2 – 85) | 1044 | WHO Encounter<br>form |
| Burns et al. 2011.<br>Causal attributions, pathway to care and clinical features of first-episode psychosis: A South African perspective | South Africa | Adolescents and<br>Adults (17 – 45) | 54 | Interview |
| Ibrahim et al. 2016<br>Pathways to psychiatric care for mental disorders: a retrospective study of patients seeking mental health services at a public psychiatric facility in Ghana | Ghana | Adults | 107 | Modified WHO<br>Encounter form |
| Lund et al. 2010<br>Pathways to inpatient mental health care among people with schizophrenia spectrum disorders in South Africa | South Africa | Adults | 152 | Interviews |
| Mkizie et al. 2004<br>Pathways to mental health care in KwaZulu – Natal | South Africa | Children and<br>Adults (10 – 59) | 15 | Interviews |
| Modiba et al. 2001<br>Profile of Community Mental Health Service Needs in the Moretele District (North-West Province) in South Africa | South Africa | Children and<br>Adults (8 – 81) | 68 | Interviews |
| Jack-Ide et al. 2013<br>Pathways to mental health care services in the Niger Delta region of Nigeria | Nigeria | Adults | 50 | Semi-structured<br>interview |
| Gureje et al. 1995<br>Pathways to psychiatric care in Ibadan, Nigeria | Nigeria | Adults | 159 | WHO Encounter<br>form |
| Gureje et al. 2006<br>Use of mental health services in a developing country. Results from the Nigerian survey of mental health and well-being | Nigeria | Adults | 4984 | Interviews |
| Erinosho et al. 1977<br>Pathways To Mental Health Delivery-Systems in Nigeria | Nigeria | Adults | 208 | Interviews |
| Galvin et al. 2023<br>Pathways to care among patients with mental illness at two psychiatric facilities in Johannesburg, South Africa | South Africa | Adults | 309 | Semi-Structure<br>Interview |
| Tomita et al. 2015<br>Duration of Untreated Psychosis and the Pathway to Care in KwaZulu-Natal, South Africa | South Africa | Adults | 57 | WHO Encounter forms |
| Odinka et al. 2014<br>Positive and negative symptoms of schizophrenia as correlates of help-seeking behaviour and the duration of untreated psychosis in south-east Nigeria | Nigeria | Adults | 360 | WHO Encounter forms |
| Bella-Awusah et al. 2020<br>Pathways to Care for Children with Mental Disorders and Epilepsy Attending Specialist Clinics in Nigeria | Nigeria | Children | 114 | WHO Encounter forms |
| Ikwuka et al. 2016<br>Pathways to mental healthcare in south-eastern Nigeria | Nigeria | Adults | 706 | Interviews |

### Prisma flow diagram

Fig 1 summarises the paper selection process using the PRISMA flow diagram. Overall, the electronic database search produced 2505 search results: 1251 from Embase, 1164 from MEDLINE, and 62 and 25 from CINAHL & WHO-Global Index Medicus, respectively. Results were then exported to Endnote, where manual screening was done for additional duplicates. After the deduplication process, 1468 papers were screened through the title and abstract. We retrieved 197 articles for full-text screening, of which twenty-seven met the inclusion criteria.

**Fig 1.**
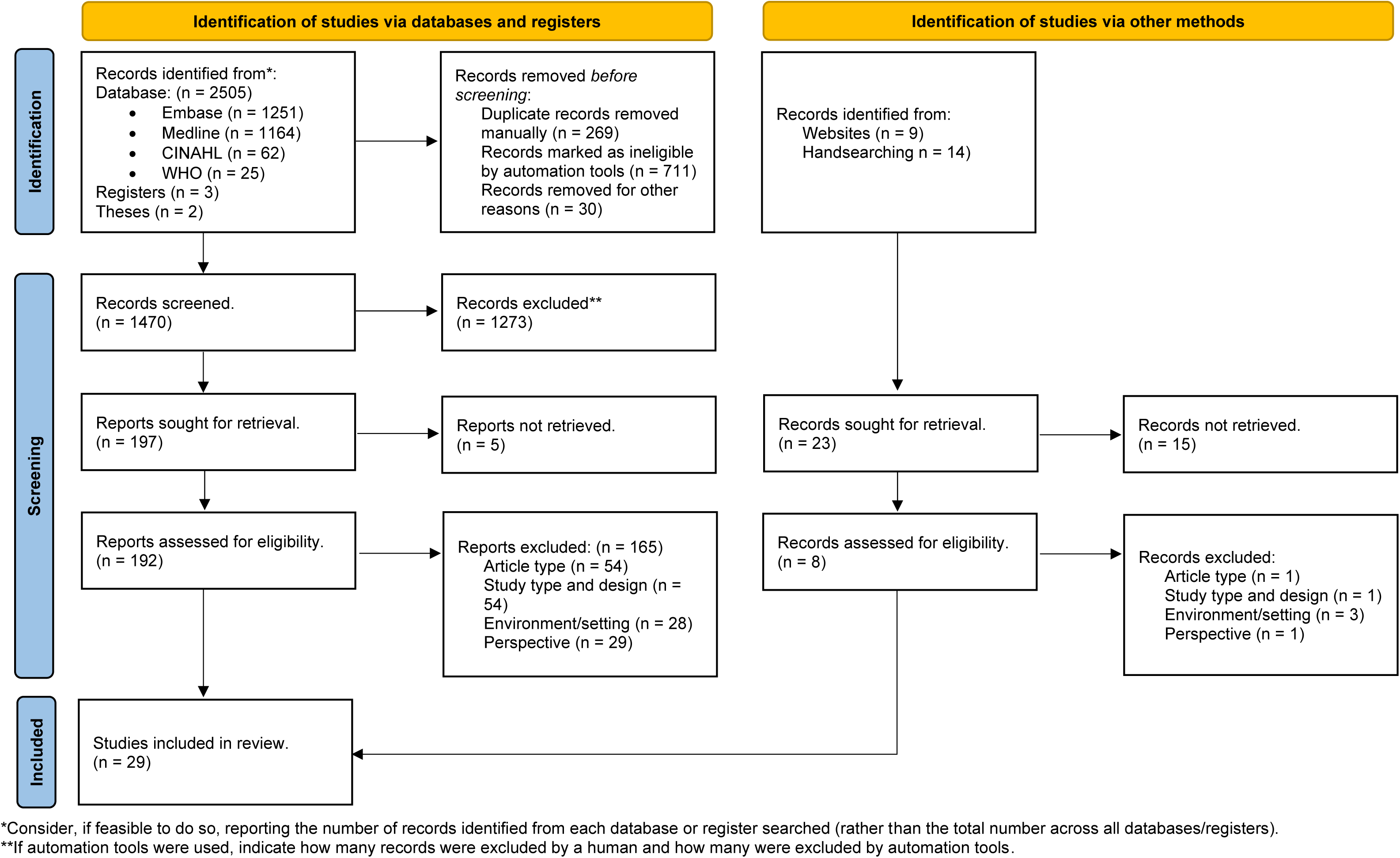
Prisma flow diagram

Additionally, through reference citations and hand searching, we identified twenty-three articles and organisational reports for screening. Eight were retrieved for full-text screening and assessed for eligibility criteria. Two additional articles met the inclusion criteria. Therefore, our final review included twenty-nine papers, four exclusively focusing on children and adolescents and twenty-five with varying populations, primarily adults. All included studies were in English.

### Chronology of included publications per year, January 1977 - September 20223

Fig 2 presents the included articles based on the inclusion criteria. All included studies were peer-reviewed articles. We identified one study from 1977, two studies from 1995 and one study from 1997, the earliest published literature in this review. Seven studies were included between 2000 and 2009, while sixteen were from the following decade, spanning 2010 to 2019. We identified and included two additional studies during the COVID-19 period spanning 2020 – 2023.

**Fig 2.**
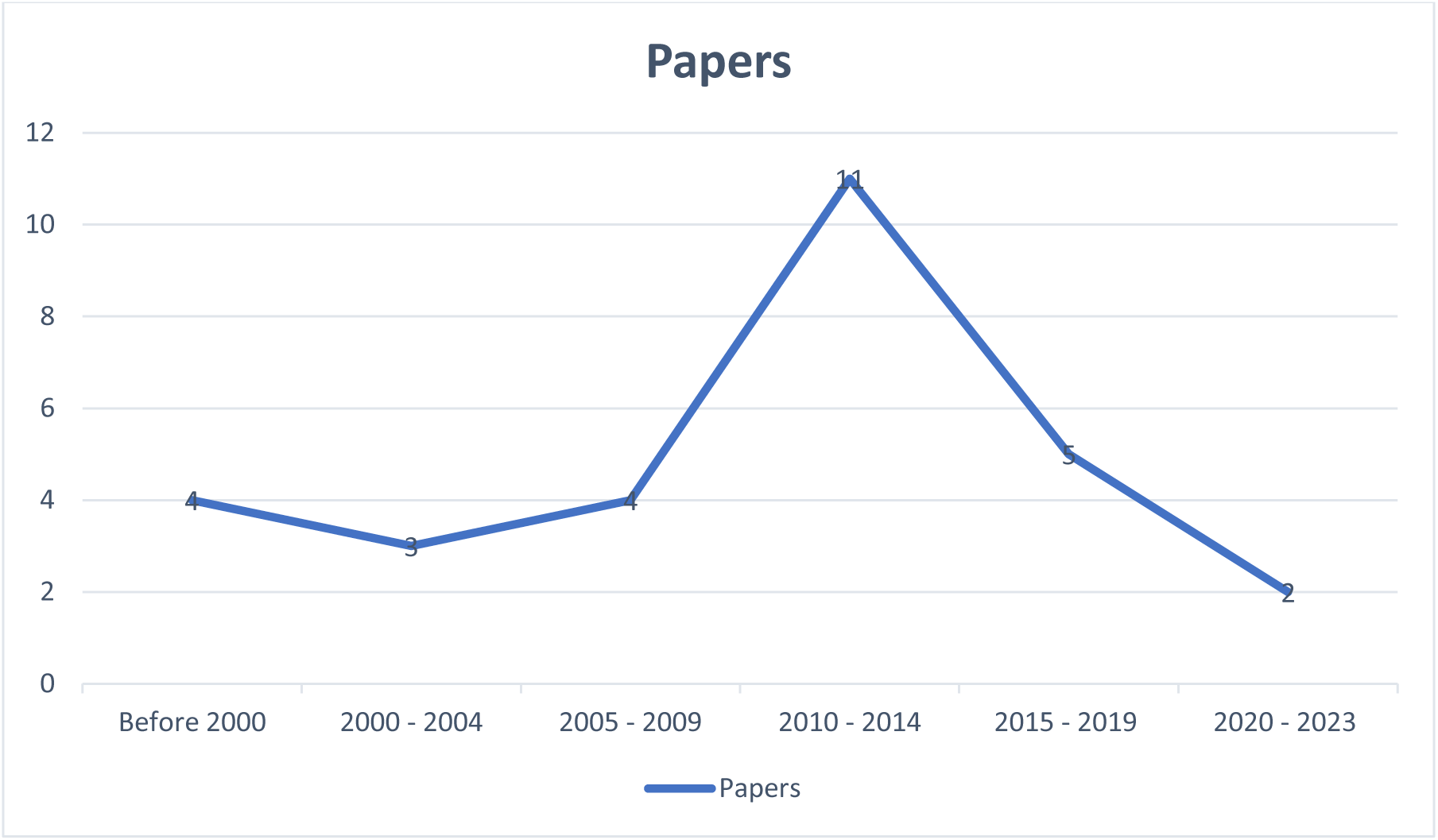
Chronology of included publications

**Fig 3.**
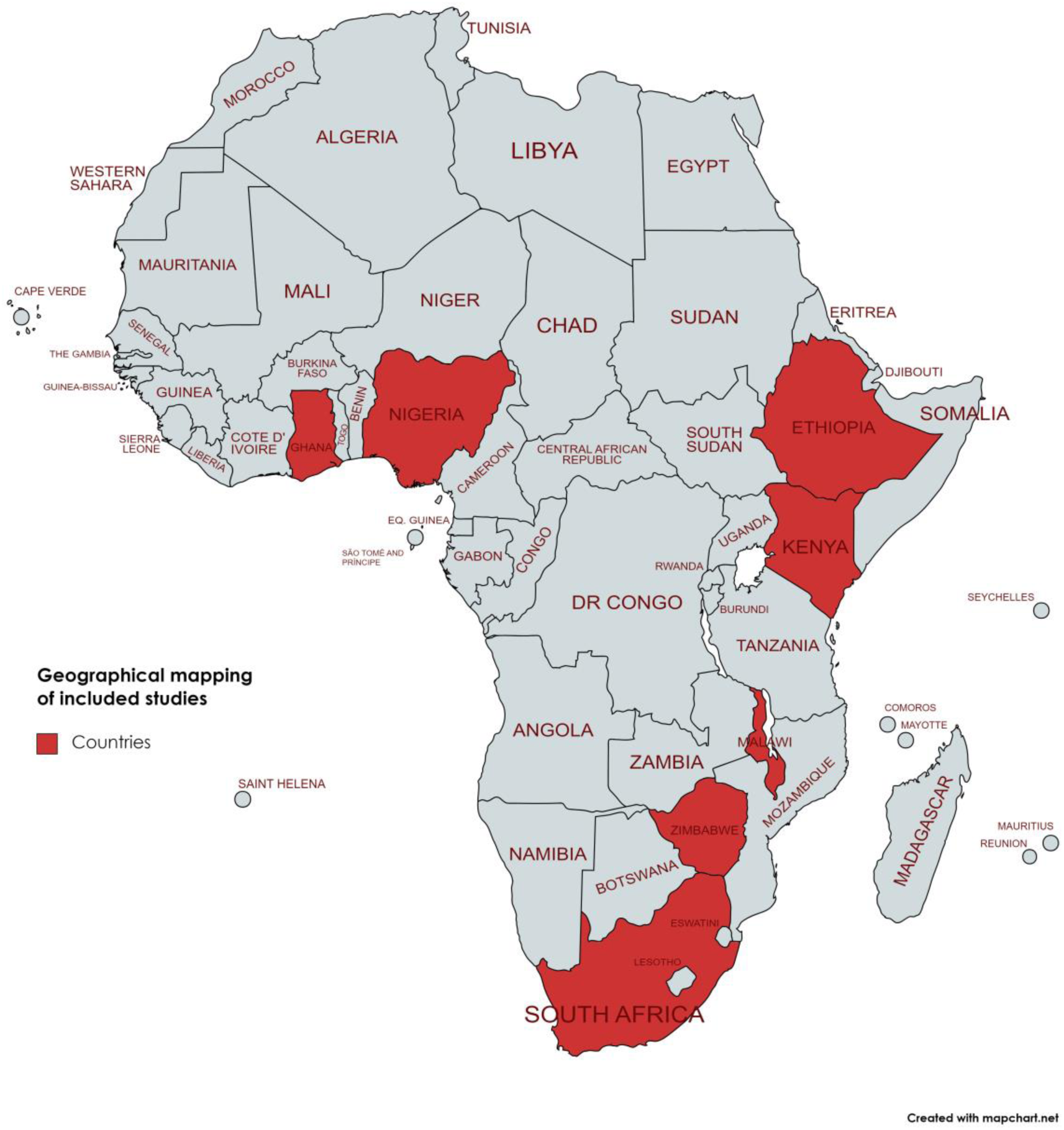
Geo-spatial locations of included studies

### Geo-spatial locations of included studies

This review draws from three of the four regions in SSA (East, South, and West Africa) and includes studies from seven countries. About 58% (17/29) of the articles/journals are from West Africa, specifically Ghana and Nigeria, with Nigeria having the bulk (fifteen) of the articles in the review.

East Africa (Ethiopia, Kenya, and Malawi) has five studies, while Southern Africa (South Africa and Zimbabwe) has eight studies. South Africa contributed eight studies, the second highest number after Nigeria. We did not find any eligible study from Central Africa. More than 70% of the studies were located in two countries, Nigeria (Abiodun, 1995; Gureje et al., 1995; Nonye & Oseloka, 2009; Abdulmalik & Sale, 2012; Aghukwa, 2012; Lasebikan et al., 2012; Jack-Ide et al., 2013; Adeosun et al., 2013; Bakere, 2013; Odinka et al., 2014; Ikwuka et al., 2016; Bella – Awusah et al., 2020; Erinosho, 1977; Gureje & Lasebikan, 2006) and South Africa (Modiba et al., 2001; Mkize & Uys, 2004; Temmingh & Oosthuizen, 2008; Burns et al., 2011; Galvin et al., 2023; Tomita et al., 2015; Lund et al., 2010). The most extensive individual study, however, is from Nigeria (Bekele et al., 2009), with a sample size of 4984 participants, representing about 39.9% of the total sample size (total participant number) of all the included studies (N= 12,491).

### Research context and approach

#### Reported study methods

Fig 4 presents the study methods employed by each article. Fifteen studies utilised various interviews (three semi-structured, eleven structured interviews, and one clinician-administered interview), nine used the WHO Encounter form, four used a modified/ adapted WHO Encounter form, and one used the Pathways to Care Schedule. Four studies focused exclusively on the pathways to care for children and adolescents: three in Nigeria (Abdulmalik et. al., 2012; Bakere, 2013; Bella-Awusah et al., 2020), and one in Kenya (Kamau et al., 2017). The other studies primarily focused on adult and caregiver populations, while some included children as young as two-year-old (Bekele et al., 2009). Burns et al. 2011 only asked if participants had sought care from traditional healers before; details of other pathways are not described.

**Fig 4.**
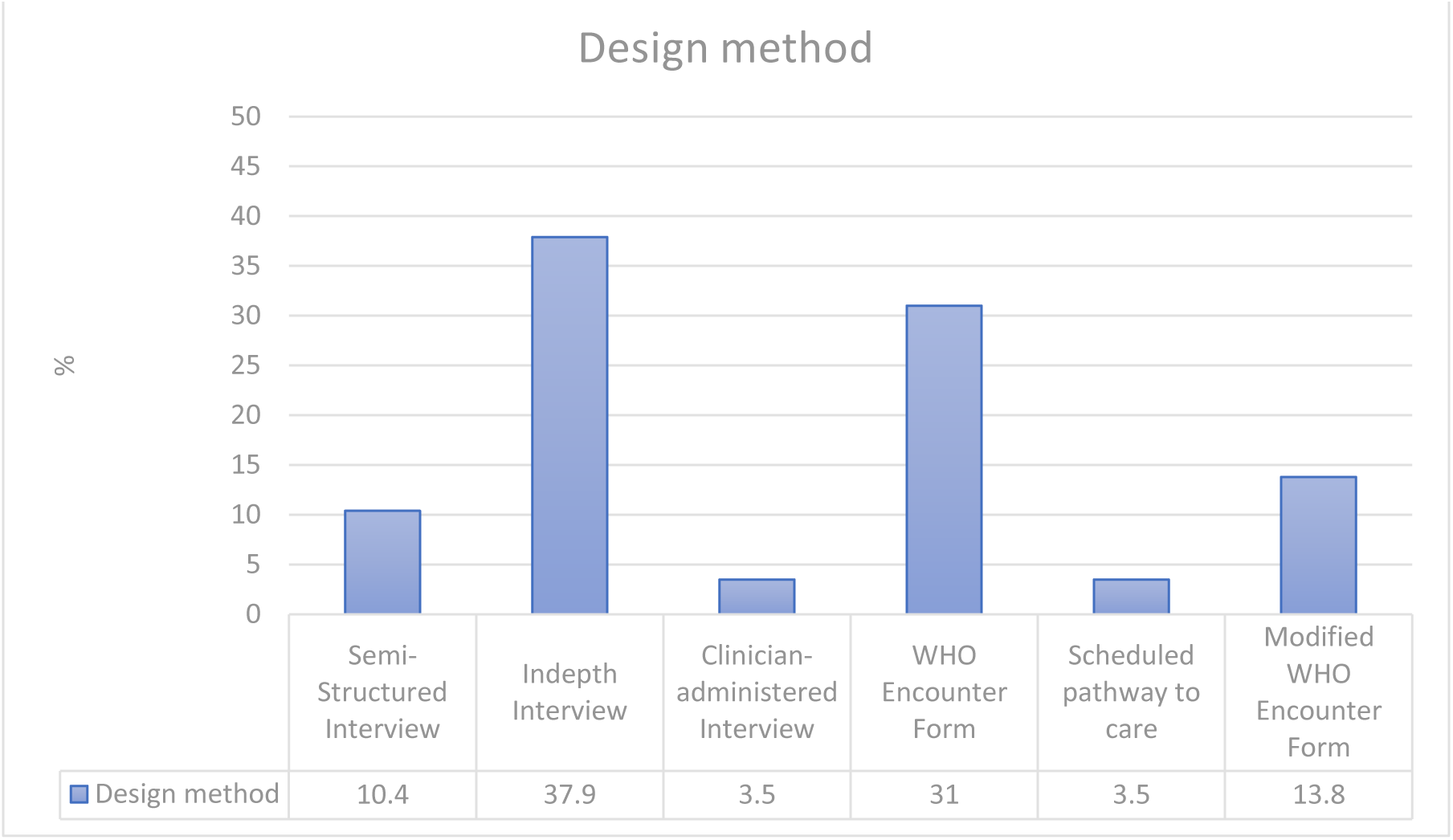
Reported study methods

### Pathways to Care

Table 2 presents various routes that patients and caregivers take to access healing services for severe mental health issues. Although many people consulted tertiary mental health services directly, the various routes can be broadly classified into formal and informal pathways.

**Table 2.** Pathways to Care

| Paper | Traditional healers (%) | Religious leaders/ healers (%) | Traditional/ Religious (undifferentiated) (%) | General practitioners (GPs) (%) | Direct tertiary (%) | Police (%) | Others (%) | Did not report (%) |
| --- | --- | --- | --- | --- | --- | --- | --- | --- |
| Abdulmalik et al. 2012 | - | - | 36.4 | 3.3 | 60.2 | - | - | - |
| Bakare et al., 2013 | 6.9 | 22.4 | - | 20.6 | 47.6 | - | Patent stores, special schools: 2.6 | - |
| Kamau 2017 | - | 6 | - | 31.9 | 52.4 | - | Pharmacy: 0.6<br>Juvenile justice system: 0.6<br>School counselor: 3.6<br>Change environment: 4.2 | - |
|  |  |  |  |  |  |  | Relatives: 0.6<br>School assessment<br>centres: 1.8 |  |
| Abiodun et al.<br>1995 | 26.5 | 13.4 | - | 55.9 | - | - | Patent medicine<br>dealers: 4.2 | - |
| Adeosun et al.<br>2013 | 11.6 | 42.8 | 14.5 | 13.8 | 17.4 | - | - | - |
| Patel et al.<br>1997 | 24.5 | 10 | - | 21.9 | - | - | Primary care<br>clinic: 42.7 | - |
| Temmingh et<br>al. 2008 | - | - | 5.6 | 39.4 | 29.6 | 25.4 | - | - |
| Odinka et al.<br>2015 | 14.7 | 61.1 | - | 14.7 | 6.4 | - | Drug store: 3.1 | - |
| Nonye et al.<br>2009 | 13.6 | 34.5 | - | 15.9 | 32 | - | Patent medicine<br>vendor: 4 | - |
| Lasebikan et<br>al. 2012 | - | - | 78.9 | 9.3% | 4.5 | - | - | - |
| Kauye et al.<br>2014 | - | - | 26.1 | 46.9 | - | 2.7 | Psych nurse: 24.3 | - |
| Girma et al.<br>2011 | 20.1 | 30.2 | - | - | 35.2 | - | Biomed Institute:<br>13.3 | - |
| Appiah-Poku<br>et al. 2003 | 5.9 | 14.2 | - | 52.8 | 2 | - | Family doctor: 4.7 | - |
| Aghukwa e al.<br>2012 | 28 | 69 | - | 24 | - | - | Other health<br>professionals: 3 | - |
| Bekele et al.<br>2008 | 4.5 | 30.9 | - | 21.5 | 41 | - | Psych nurses: 2 | - |
| Burns et al.<br>2011. | 38.5 | - | - | 16 | - | 44 | - | - |
| Ibrahim et al.<br>2016 | - | - | 23.3 | 21.5 | 52.3 | - | Community health<br>nurse: 2.9 | - |
| Lund et al.<br>2010 | - | - | - | 73 | 29 | 51 | - | - |
| Mkizie et al. | 20 | 20 | - | 33.3 | - | - | District Hospital: | - |
| 2004 |  |  |  |  |  |  | 20<br>Primary<br>Healthcare clinic:<br>6.7 |  |
| Modiba et al.<br>2001 | 30 | - |  | 70 | - | - | Friends: 7 | - |
| Jack-Ide et al.<br>2013 | 20 | 48 | - | 20 | 12 | - | - | - |
| Gureje et al.<br>1995 | 19 | 13 | - | 47 | 20 | - | - | - |
| Gureje et al.<br>2006 | - | - | 3.7 | 28.2 | 29.1 | - | 32.8 | - |
| Erinosho et al.<br>1977 | 0.5 | - | - | 4.3 | 3.4 | - | Family: 89.4 | - |
| Galvin et al.<br>2023 | 28.16 | 15.04 | 4.7 | 53 | - | - | - | - |
| Tomita et al. | - | - | 11.5 | 3.9 | 73.1 | 1.9 | Clinic: 3.9 | - |
| 2015 |  |  |  |  |  |  | Social Worker: 1.9<br>Private<br>Psychiatrist: 3.9 |  |
| Odinka et al.<br>2014 | 14.7 | 61.1 | - | 17.8 | 6.4 | - | - | - |
| Bella-Awusah<br>et al. 2020 | - | 54.39 | - | 31.58 | 14.04 | - | - | - |
| Ikwuka et al.<br>2016 | 33.2 | 57.8 | - | 90.8 | - | - | - | - |

This review identified formal pathways as general practitioners (GPs), primary health care clinics, general and regional hospitals, paramedics, nurses, pharmacists, drug stores, psychiatrists, and tertiary mental health hospitals. Informal routes include traditional healers, religious healers, and patent medicine vendors. Specialised schools and the juvenile justice system were also used as a pathway to care for children and adolescents (Bakele et al., 2013; Kamau et al., 2017).

### Pathways to Care for Children and Adolescent

This review finds that the most frequently used care pathway by caregivers for children and adolescents was the direct route to tertiary hospitals, followed by general health services and practitioners. It supposes that biomedical treatment is the preferred option for children and adolescents.

The estimate for caregivers who seek mental health services directly from tertiary care ranges from 47.6% to 60.2% from three studies on children and adolescents (Abdulmalik et al., 2012; Bakere, 2013; Kamau et al., 2017). This is followed by those seeking care in general medical services, with an estimated 20.6% to 37.9% (Bakere, 2013; Kamau et al., 2017). Only 3.3% of caregivers sought care from general practitioners in Nigeria (Abdulmalik et al., 2012).

A considerable number of people, however, consulted traditional or religious healers as their first choice of care. Results from these four studies on children revealed an average of 29.3% to 47.1% of caregivers consulted traditional or religious healers as their primary source of care, except in the study by Kamau et al. 2017, where only 6% of patients or caregivers contacted religious leaders.

Their study nevertheless identified other varied and important routes taken by patients/ caregivers in need of help. These routes include consulting school counsellors, accessing the juvenile justice system, and seeking pharmacy help.

### Pathways to care for adults

Twenty-five studies were analysed for pathways to care in the adult population. This review shows that general medical services, tertiary hospitals, and traditional/ religious healers were the main pathways to care used by patients with mental health disorders.

Patients in ten of these studies preferred biomedical health services for treatment modalities (Abiodun, 1995; Gureje et al., 1995; Patel et al., 1997; Modiba et al., 2001; Appiah-Poku et al., 2004; Mkize & Uys, 2004; Temmingh & Oosthuizen, 2008; Bekele et al., 2009; Kauye et al., 2015; Ibrahim et al., 2016).

Six of the eight studies that showed a preference for traditional and religious healers were conducted in Nigeria over the last fifteen years, with the earliest published in 2009. In contrast, the other two that preferred biomedical services were conducted in 1995.

In the studies that found a preference for traditional/ religious healers, this evaluation found averages as high as 78.9% of the population consulting traditional/ religious healers as the first source of care (Lasebikan et al., 2012). Conversely, in Temmingh & Oosthuizen, 2008 study, only 5.6% of participants consulted them. The highest average for patients seeking biomedical care ranges between 73.8% and 75.5% (Patel et al., 1997; Ibrahim et al., 2016). In two other studies, the researchers sought only to quantify the number of people who have sought care from traditional healers.

In another study by Burns et al. 2011 in Kwa-Zulu Natal, South Africa, 38.5% of patients admitted at the tertiary hospital sought care from traditional healers as the first point of care, with only 35% seeking care from biomedical services (19% from psychiatrists).

This review, however, notes that many people used several other help-seeking pathways for mental health service, including the police (2.9% to 25.4%) (Temmingh & Oosthuizen, 2008; Modiba, 2001; Kauye et al., 2015), nurses (2% to 21.1%) (Bekele et al., 2009; Kauye et al., 2015), patent medicine vendors/ pharmacies (3.1% to 4.2%) and other community health workers (Abiodun, 1995; Nonye et al., 2009; Bakere et al., 2013; Kamau et al., 2017).

### Socio-demographic correlates of Pathways to care

#### Studies on children and adolescent

In the five studies exclusively on pathways to care, female gender, level of education, age of patients, social and psychological barriers, and being accompanied by both parents were the only significant socio-demographic factors. Patients (children) whom both parents accompanied to the psychiatric facility were found to be two and a half (OR=2.5, 95% CI 2.35-2.6) times more likely to have had a mental illness for less than six months compared to patients who were single parents accompanied. In contrast, caregivers’ education and social status did not correlate significantly with the care pathway (Abdulmalik, 2012).

Female gender and patients without disruptive disorders were the factors that were significantly associated with the pathway choice in the study by Kamau, 2017. Females were significantly more likely to be taken through the biomedical pathway by caregivers than males (p=0.027), and this was the same for participants without disruptive behaviours compared to those with disruptive behaviours compared to their counterparts.

#### Studies on adults

Out of the twenty-five eligible studies, only eleven found at least one factor to be significantly associated with the choice of pathway. These factors include age, sex, marital status, education, geographic location (rural or urban), knowledge about aetiology, family contact or support, employment, and aggressive behaviours.

In the study by Odinka et al. 2014, participants who were below the age of 40 years were significantly more likely to use religious faith healers as their first option for a pathway to care when compared to those above 40 years who preferred traditional or faith-based healers as their first option (Odinka et al., 2014). Another study found participants who were between the ages of 31 and 40 years to be more than ten times less likely to seek treatment when compared to those below the age of twenty (OR-10.7, 95% CI 1.99-56.99) (Girma & Tesfaye, 2011).

Another important correlate was education; patients who have spent more than six years in education were more likely to use Western medicine than those with fewer years in education (Odinka et al., 2014). This was similar to the findings by Nonye et al. 2009, who established that a higher level of education was statistically significantly associated with specialist service usage as their first pathway to care. Patients who lacked formal education experienced a significant delay from starting symptoms to reaching the hospital for mental health treatment (Bekele et al., 2009). In contrast, patients with secondary school education were less likely to seek care from specialist psychiatric public hospitals when compared to patients with no formal education (uOR = 0.86; 95 % CI 0.18– 4.08) (Ibrahim et al., 2016).

In terms of location, patients living in rural communities were found to be more likely to use traditional medicine when compared to people from urban communities (Odinka et al., 2014); this was also supported by Nonye et al. 2009, where he made a statistically significant observation that urban community dwellers are more likely to use specialist mental health service compared to those in rural communities. Attribution of the causation of mental illnesses also played a role in the care pathway. Correct rationalisation of the cause of the mental illness was associated with consultation with specialist medical services (Nonye & Oseloka, 2009). In contrast, spiritual and/or traditional attribution of causation and previously consulting traditional healers were associated significantly with a long delay of untreated psychosis (DUP) (Burns et al., 2011).

Regarding employment status, self-employed patients had almost twice the odds of seeking mental health care in general medical health facilities and more than four times higher odds (OR-4.4) with public servants than unemployed patients (Ibrahim et al., 2016). People who were unemployed/ jobless were also found to experience significantly longer times (delays) in accessing mental health care from the onset of symptoms to the first point in the care pathway and then reaching a formal psychiatric service (Bekele et al., 2009). Other important correlates that were significant in determining access to formal mental health services and early treatment seeking were male gender (Nonye & Oseloka, 2009), a family member with psychiatric illness, marital status, presence of somatic symptoms (Girma & Tesfaye, 2011), and those in full contact with family almost every day (Lasebikan et al., 2012).

#### Duration/ delay in seeking treatment

The distribution measurement for the delay in seeking care was highly skewed in most reported studies; therefore, the median was used. For children and adolescents, the median delay from the onset of symptoms to accessing care at a formal health service was between 4.5 and 54 months [inter quartile range (IQR)=22.6 months] (Kamau et al., 2017). In another study, 64.5% of patients at a psychiatric hospital in Nigeria were found to have been with a mental illness for more than six months before presenting to the hospital (Abdulmalik, 2012).

Evidence from studies on the adult population indicates that the overall median delay from symptom onset to presentation at the hospital was between 4.5 and 9.5 months (Temmingh & Oosthuizen, 2008; Bekele et al., 2009; Appiah-Poku et al., 2004), with an IQR between 35 to 37 months.

Aghukwa et al. 2012 reported a mean of 54 months from disease onset to psychiatric consultation in the hospital. When the different pathways that were sought were factored in, a patient who first consulted traditional or faith-based healers were found to have experienced the most extended delay when they arrived at the hospital compared to those who consulted formal health services directly, and this association was significant (Abiodun, 1995; Gureje et al., 1995; Appiah-Poku et al., 2004; Bekele et al., 2009; Adeosun et al., 2013; Odinka et al., 2014; Kauye et al., 2015).

### Patients/caregivers’ perception of the cause of mental illness and sources of referral

The perception of causes of mental illness from the reviewed literature can be broadly classified into spiritual, environmental, and medical causes, with some participants admitting that they do not know. Spiritual belief emerged as one of the main reasons patients consulted with traditional and religious healers. In a study conducted in Nigeria, 71% to 85% of patients consulted with and preferred traditional/ religious healers because they believed in the supernatural origin of the illness (Aghukwa, 2012; Adeosun et al., 2013), and this was reported by only 3.2% of patients in Zimbabwe (Patel et al., 1997). On average, 29.1% to 69.4% of patients (caregivers) believed that mental illness was due to spiritual causes (Nonye & Oseloka, 2009; Burns et al., 2011; Girma & Tesfaye, 2011; Aghukwa, 2012).

The evidence also suggests that only 22.3% to 41% of patients in the reviewed studies believe in the biomedical causation of mental illness (Aghukwa, 2012; Burns et al., 2011; Nonye & Oseloka, 2009). Between 19.6% to 48.6% of participants acknowledge they have no idea about the causes of mental illness (Girma & Tesfaye, 2011; Nonye & Oseloka, 2009), while 16.9% associated them with environmental factors (Aghukwa, 2012).

## Discussions

This study reviewed the evidence in the literature on the pathways to mental health care in SSA. It explored the socio-demographic correlates that affect help-seeking behaviour and delay in seeking care (Fig 5).

**Fig 5.**
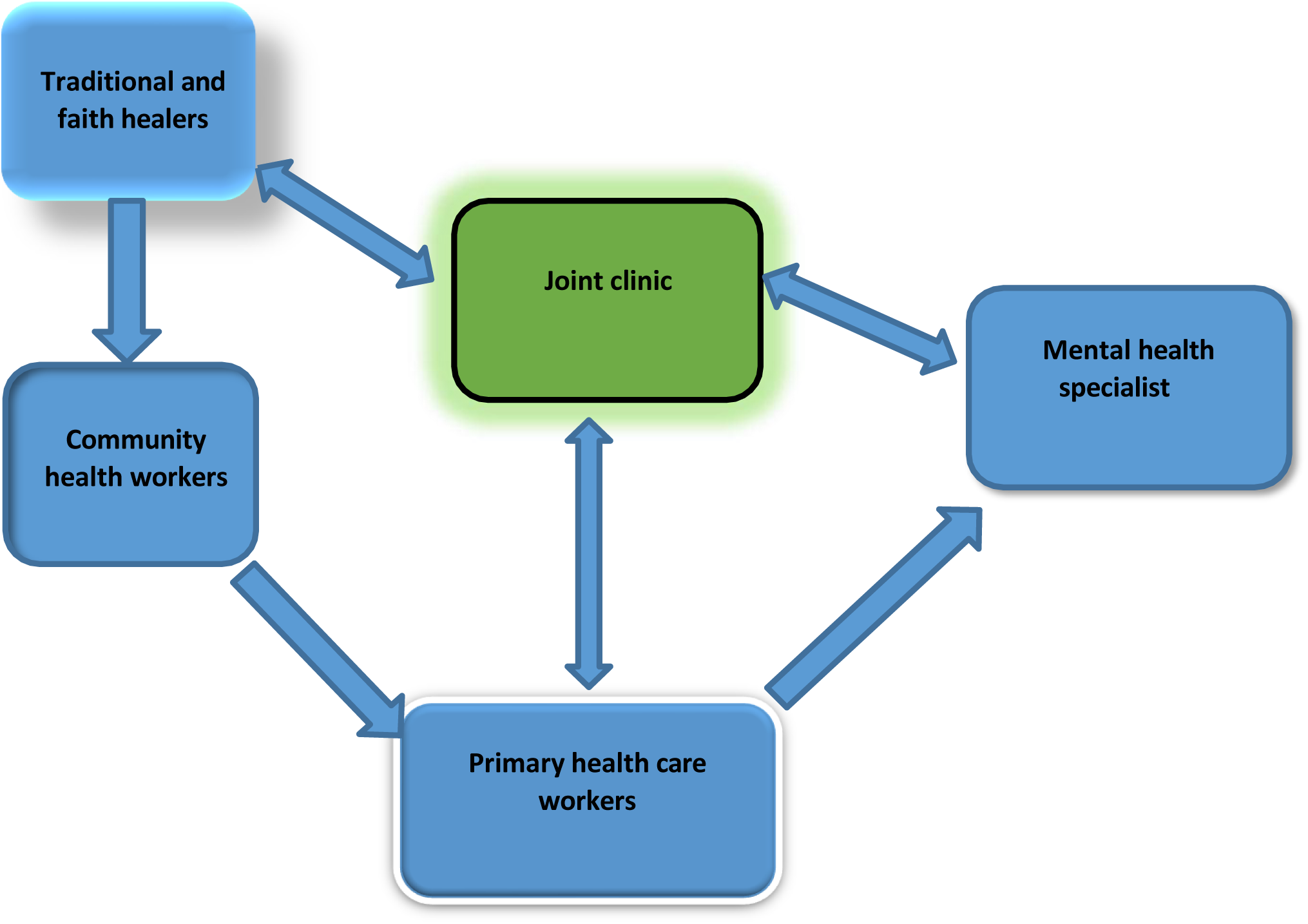
Referral Pathway

The result mainly reflects medical pluralism as reported for different health issues in Africa. The fact that a tertiary level is directly scarcely consulted may be related to the fact there are very few psychiatrists in these countries and are mainly located in big cities and tertiary hospitals. This reflects the need to decentralize MH services. Patients and caregivers use a combination of biomedical and traditional or faith-based pathways to mental health care and strongly suggest a model in the SSA context to enhance early detection to improve prognosis. The synthesised studies showed that many patients who presented for mental health care at formal services had consulted with traditional or faith-based healers as the first point of care. In one of these studies, it was observed that up to 78.9% of the patients had consulted traditional or faith-based healers as the first point of care. This finding is similar to that reported by the WHO and other previous studies (mhGAP WHO, 2008; Mbwayo et al., 2013).

However, the evidence indicates that most of the articles in the review showed an overall preference for biomedical health services as a preferred treatment option, although it also varied with education, age, and gender. In contrast to the study by Badu et al. 2019, many patients who visit these formal services directly visit tertiary mental health facilities or psychiatrists as the first point of care without any referral from other services. One of the articles from Nigeria revealed that as many as 60.2% of patients or their caregivers directly sought help from tertiary services.

Six of the eight articles that showed an overall preference for traditional or faith-based healers were from Nigeria, with one each from Ethiopia and South Africa, respectively. This finding is similar to that of Burns et al. 2015 in a systematic literature review. This occurrence might, therefore, significantly influence the generalizability of this finding to the African continent, given the predominance of the result from a single country, as it might lead to an ’over-estimation’ of the proportion who sought mental health care from traditional or faith-based healers. This result should, however, consider specific cultural factors or causal attributions that might influence this preference in the pathway to care.

However, recursive pathways exist where patients move between formal and informal services (Mkize et al., 2004; Bekele et al., 2009; Girma & Tesfaye., 2011; Adeosun et al., 2013; Kauye et al., 2015), consistent with findings from other LMICs, including Africa (Badu et al., 2019; Burns & Tomita, 2015; Giasuddin et al., 2012).

### Delay in access to care

As alluded to earlier, one of the most important aspects in considering help-seeking behaviour must be how the choice of first care provider for mental health affects or leads to delay in accessing appropriate treatment. Delay in treatment result in increased morbidity and mortality, including significant harmful health effects to both the society and individual (e.g., substance abuse, psychiatric commodities, and life-altering self-treatments).

This review found that patients who sought care from traditional and faith-based healers as the first point of care were found to have experienced the most prolonged delay in accessing evidence-based mental health services at tertiary hospitals. Complicated patterns of correlation indicate multiple causative factors for delay, such as educational attainment, income status, availability of healthcare services, and cultural barriers. The lack of knowledge regarding MH is a major concern that requires adequate assessment and action plan. Strategies that reduce delays in accessing mental healthcare services and early interventions in patients with severe mental illnesses will decrease the morbidity associated with the illness (O’Callaghan et al., 2010; Morgan et al., 2006).

This delay in care shown in this study should inform strategies on how formal biomedical services and policymakers/planners should engage with traditional or faith-based healers and other informal health providers in Africa to improve the pathways to care for persons with mental illnesses. Even though the aim is to shorten the delay by getting patients with mental health issues into formal health services quickly, it is difficult to achieve this in these settings where resources are limited, and structures or services to aid this process are not readily available in the communities (Burns & Tomita, 2015). Inadvertently, the mental health needs of the patients, caregivers, and communities are unmet by the existing health policies and services (Burns, 2014).

### Collaborative model

#### Role of Stakeholders/Key Players

This review has identified that traditional or faith-based healers play a significant part in the mental health care pathway in Africa, particularly in Western Africa, and much research has shown the need for collaboration between formal and non-formal services (Ae-Ngibise et al., 2010). This review proposes that the pathway to care for mental illnesses would benefit from similar models used in other public health programs, such as the TB and HIV programs that have successfully implemented collaborative care models with traditional or faith-based healers in some African countries (Peltzer et al., 2006; Kayombo et al., 2007; Harper et al., 2004; Audet et al., 2013). This aligns with the recommendations by Patel, 2011, who described this as a rewarding partnership to reduce the treatment gap in Africa and should be the objective for global mental health. Nonetheless, several documented barriers have prevented such collaboration. These factors are pervasive in all sectors of mental health care delivery.

Stakeholders in formal care pathways have raised concerns about the safety and efficacy of interventions employed by traditional or faith-based healers and their efficiency and reports of human rights abuses against persons with mental illness in these stings (Read et al., 2009; WHO, 2013). It should also be noted that cases of abuse are not peculiar to TAFH, as there have been harrowing reports of human rights abuses even in tertiary mental health facilities (Kelly, 2006; WHO, 2018).

On the other hand, traditional healers have also expressed concerns about working with healthcare workers. These stem from the fear of reproducing their treatment techniques and concoctions in scientific labs (Kilonzo & Simmons, 1998; Wreford, 2005). Another concern is that Western medicine is superior to traditional healing techniques, and the latter should not be recognised (Van der Watt et al., 2017)

Despite all the doubts expressed on both sides of the healing services, some traditional or faith-based healers and healthcare professionals have signaled the willingness to work together (Ae-Ngibise et al., 2010) when mutual respect is achieved through understanding and appreciating local beliefs and practices.

Therefore, implementing a proposed collaborative model cannot be thoroughly carried out if these concerns are not adequately addressed. A collaborative model’s goal is to foster a working relationship between the two care systems and implement a clear referral pathway. It will also serve as a platform where the reasons for ’unsafe’ interventions are investigated and researched, with proper solutions and behavioural change implemented. This model aims to put the needs and welfare of the patient at the centre, where there is a shared responsibility between the providers they choose to consult, which is facilitated by mutual respect and understanding. Formal mental health services can also use this opportunity to learn from traditional or faith-based healers’ psychosocial approach, given that the main goal is to harness treatment modalities that can promote the health of patients with mental health problems. The model does not attempt to interfere with the duties of both traditional and biomedical services but rather train traditional or faith-based healers on evidenced-based psychosocial interventions and open referral pathways between the two services.

The WHO Mental Health Global Action Programme intervention guide (MhGAP-IG) intervention guide was designed to be used by non-specialists in health facilities at primary care levels. It recognises traditional and faith healers as non-specialists and a potential resource to reduce the treatment gap in low- and middle-income countries (Mental Health Atlas, 2020; Oduguwa et al., 2017). However, MhGAP must be adapted to suit their local context, resources, priorities, and other mental illnesses not initially included. Adapting and modifying the MhGAP based on the results of context-specific studies will facilitate an effective interventional approach.

A collaborative model herein will enhance the easy and quick identification of mental health cases at the community level by CHWs and traditional and religious healers. The new model will also enhance and build upon weak referral pathways by collaborating with these community members to identify persons with mental health problems, offer appropriate evidence-based psychosocial interventions, and refer complex cases directly to community primary care clinics or tertiary hospitals.

## Strengths and limitations

This review has shown that traditional or faith-based healers play an integral role in the pathways to care in African countries. Given the burden of mental illnesses in SSA and the lack of resources for mental health, there is an urgent need to mobilise available resources for managing mental health illnesses. Our results have the potential to shape standards for health service delivery involving mental health care and enhance the optimisation of access on a regional scale. The findings may interest a wide range of stakeholders, including researchers, funders, and policymakers, all aiming to comprehend the fundamental concepts behind increased access to mental health services. Additionally, there is a possibility that patients and their carers, being central stakeholders, could benefit to some extent. Increased awareness and knowledge could enable decision-makers to contribute to developing strategies that promote a smoother policy and intervention integration.

Our review could be influenced by language and reporting biases. The studies in our analysis presented results of cross-sectional help-seeking behaviour using standardised questionnaires (WHO Encounter form and Pathways to Care schedule) to assess the pathways to care. Consequently, our systematic review likely provides only a glimpse of the real-world barriers, inevitably overlooking some challenges to access and policy implications in practice. Therefore, we acknowledge that our analysis may represent a scenario where only a fraction of the actual pathway and access to mental healthcare is revealed, akin to the metaphorical “tip of the iceberg.”

Another important limitation stems from the fact that almost all the studies in this review were conducted at a tertiary mental health facility, with a few conducted in other formal health services such as clinics. Many people suffering from mental health disorders who have never had access to formal services or have never attempted to consult with them might have a different help-seeking behaviour not reflected in the results below. Thus, it cannot be said that the result represents all African populations that require or seek help for mental health problems, mainly because formal health services are notoriously few and often inaccessible to remote communities.

## Conclusion

This review proposes and recommends a new model for collaboration between biomedical and traditional or faith-based healers that focuses on education through training and adopting a new referral framework premised on mutual respect and appropriate incentives. Therefore, the collaborative model’s goal should be to harness all the practices of traditional or faith-based healers that promote health to benefit the mentally ill. It is also important to note that the focus on improved access to mental health care in Africa should focus on other innovative strategies if the large treatment gap is to be reduced due to the chronic lack of human and structural resources. One such strategy, in addition to collaboration, should also focus on fully integrating mental health care into general and primary health care settings, with appropriate training of health workers even at the community level.

## Data Availability

All data produced in the present work are contained in the manuscript

https://www.crd.york.ac.uk/prospero/display_record.php?ID=CRD42023459738

