## Supporting Information for "Pathways to mental health services across local health systems in sub-Saharan Africa: Findings from a Systematic Review"

### S1. Search Strategies Legend

We created specific search strategies for each database using index terms related to 'pathways to care,' 'mental health,' and 'Africa'. We took cues from the following research to guide the development of our search strategy.

For ‘pathways to care’ search terms:

Daghash, H., Abdullah, K.L. and Bin Ismail, M.D., 2019. The strategy of development and implementation of care pathway: literature review. International Journal of Integrated Care, 19(4), p.569.DOI: https://doi.org/10.5334/ijic.s3569. Wilczynski NL, Marks S, Haynes RB. (2007) Search Strategies for Identifying Qualitative Studies in CINAHL. Qualitative Health Research; 17(5):705-710. doi:10.1177/1049732306294515

Latina R, Salomone K, D’Angelo D, Coclite D, Castellini G, Gianola S, Fauci A, Napoletano A, Iacorossi L, Iannone P. Towards a New System for the Assessment of the Quality in Care Pathways: An Overview of Systematic Reviews. International Journal of Environmental Research and Public Health. 2020; 17(22):8634. https://doi.org/10.3390/ijerph17228634

For ‘mental health’ search terms:

Mansfield, R., Patalay, P. & Humphrey, N. A systematic literature review of existing conceptualisation and measurement of mental health literacy in adolescent research: current challenges and inconsistencies. BMC Public Health 20, 607 (2020). https://doi.org/10.1186/s12889-020-08734-1

For ‘Sub-Saharan Africa’ search terms:

Awini E, Agyepong IA, Owiredu D, et alBurden of mental health problems among pregnant and postpartum women in sub-Saharan Africa: systematic review and meta-analysis protocolBMJ Open 2023;13:e069545. doi: 10.1136/bmjopen-2022-069545

We used the World Bank country classification 2019 (on Embase) and 2021 (on Medline) for this review and ran an expert group database search for Sub-Sahara African countries.

### S2. Table. Embase and Medline

| | |
|---|---|
| 1 | exp clinical pathway/ or pathway* to care.mp. or exp patient care/ |
| 2 | health seeking behavior/ |
| 3 | access to care/ |
| 4 | ((pathway* to care or clinical pathway* or (health adj2 behavio?r) or access to care or patient journey or healthcare pathway* or service access or care continuum or care routes or treatment) adj2 access).mp. |
| 5 | 1 or 2 or 3 or 4 |
| 6 | mental health/ |
| 7 | mental disorder/ |
| 8 | psychiatry/ |
| 9 | (mental health or (mental adj2 wellness) or mental disorder* or psychological health or emotional health or mental fitness or psychosocial well-being or behavio* health or psychiatr* wellness or Lunac* or melancholia or hyster* or nervous breakdown or madness or moral insanity or bipolar disorder* or psychotic disorder or anxiety or anxiety disorder* or depression or schizophreni*).mp. [mp=title, abstract, heading word, drug trade name, original title, device manufacturer, drug manufacturer, device trade name, keyword heading word, floating subheading word, candidate term word] |
| 10 | 6 or 7 or 8 or 9 |
| 11 | "Africa south of the Sahara"/ |
| 12 | ("Africa South of the Sahara" or sub-Saharan Africa or subSaharan Africa).ti,ab. |
| 13 | Central Africa.ti,ab. |
| 14 | Eastern Africa.ti,ab. |

| | |
|---|---|
| 15 | Southern Africa.ti,ab. |
| 16 | Western Africa.ti,ab. |
| 17 | Seychelles/ |
| 18 | Seychelles.ti,ab. |
| 19 | Benin/ |
| 20 | (Benin or Dahomey).ti,ab. |
| 21 | Burkina Faso/ |
| 22 | (Burkina Faso or Burkina Fasso or Upper Volta).ti,ab. |
| 23 | Burundi/ |
| 24 | (Burundi or Ruanda-Urundi).ti,ab. |
| 25 | Central African Republic/ |
| 26 | (Central African Republic or Ubangi-Shari).ti,ab. |
| 27 | Chad/ |
| 28 | Chad.ti,ab. |
| 29 | Democratic Republic Congo/ |
| 30 | (((Democratic Republic or DR) adj2 Congo) or Congo-Kinshasa or Belgian Congo or Zaire or Congo Free State).ti,ab. |
| 31 | Eritrea/ |
| 32 | Eritrea.ti,ab. |
| 33 | Ethiopia/ |
| 34 | (Ethiopia or Abyssinia).ti,ab. |
| 35 | Gambia/ |
| 36 | Gambia.ti,ab. |

| | |
|---|---|
| 37 | Guinea/ |
| 38 | (Guinea not (New Guinea or Guinea Pig* or Guinea Fowl or Guinea-Bissau or Portuguese Guinea or Equatorial Guinea)).ti,ab. |
| 39 | Guinea-Bissau/ |
| 40 | (Guinea-Bissau or Portuguese Guinea).ti,ab. |
| 41 | Liberia/ |
| 42 | Liberia.ti,ab. |
| 43 | Madagascar/ |
| 44 | (Madagascar or Malagasy Republic).ti,ab. |
| 45 | Malawi/ |
| 46 | (Malawi or Nyasaland).ti,ab. |
| 47 | Mali/ |
| 48 | Mali.ti,ab. |
| 49 | Mozambique/ |
| 50 | (Mozambique or Mocambique or Portuguese East Africa).ti,ab. |
| 51 | Niger/ |
| 52 | (Niger not (Aspergillus or Peptococcus or Schizothorax or Cruciferae or Gobius or Lasius or Agelastes or Melanosuchus or radish or Parastromateus or Orius or Apergillus or Parastromateus or Stomoxys)).ti,ab. |
| 53 | Rwanda/ |
| 54 | (Rwanda or Ruanda).ti,ab. |
| 55 | Sierra Leone/ |
| 56 | (Sierra Leone or Salone).ti,ab. |

| | |
|---|---|
| 57 | Somalia/ |
| 58 | (Somalia or Somaliland).ti,ab. |
| 59 | south sudan/ |
| 60 | South Sudan.ti,ab. |
| 61 | Tanzania/ |
| 62 | (Tanzania or Tanganyika or Zanzibar).ti,ab. |
| 63 | Togo/ |
| 64 | (Togo or Togolese Republic or Togoland).ti,ab. |
| 65 | Uganda/ |
| 66 | Uganda.ti,ab. |
| 67 | Angola/ |
| 68 | Angola.ti,ab. |
| 69 | Cameroon/ |
| 70 | (Cameroon or Kamerun or Cameroun).ti,ab. |
| 71 | Cape Verde/ |
| 72 | (Cape Verde or Cabo Verde).ti,ab. |
| 73 | Comoros/ |
| 74 | (Comoros or Glorioso Islands or Mayotte).ti,ab. |
| 75 | Congo/ |
| 76 | (Congo not ((Democratic Republic adj3 Congo) or congo red or crimean-congo)).ti,ab. |
| 77 | Cote d'Ivoire/ |
| 78 | (Cote d'Ivoire or Cote dIvoire or Ivory Coast).ti,ab. |
| 79 | eswatini/ |

| | |
|---|---|
| 80 | (eSwatini or Swaziland).ti,ab. |
| 81 | Ghana/ |
| 82 | (Ghana or Gold Coast).ti,ab. |
| 83 | Kenya/ |
| 84 | (Kenya or East Africa Protectorate).ti,ab. |
| 85 | Lesotho/ |
| 86 | (Lesotho or Basutoland).ti,ab. |
| 87 | Mauritania/ |
| 88 | Mauritania.ti,ab. |
| 89 | Nigeria/ |
| 90 | Nigeria.ti,ab. |
| 91 | "sao tome and principe"/ |
| 92 | (Sao Tome adj2 Principe).ti,ab. |
| 93 | Senegal/ |
| 94 | Senegal.ti,ab. |
| 95 | Sudan/ |
| 96 | (Sudan not South Sudan).ti,ab. |
| 97 | Zambia/ |
| 98 | (Zambia or Northern Rhodesia).ti,ab. |
| 99 | Zimbabwe/ |
| 100 | (Zimbabwe or Southern Rhodesia).ti,ab. |
| 101 | Botswana/ |
| 102 | (Botswana or Bechuanaland or Kalahari).ti,ab. |

| | |
|---|---|
| 103 | Equatorial Guinea/ |
| 104 | (Equatorial Guinea or Spanish Guinea).ti,ab. |
| 105 | Gabon/ |
| 106 | (Gabon or Gabonese Republic).ti,ab. |
| 107 | Mauritius/ |
| 108 | (Mauritius or Agalega Islands).ti,ab. |
| 109 | Namibia/ |
| 110 | (Namibia or German South West Africa).ti,ab. |
| 111 | South Africa/ |
| 112 | (South Africa or Cape Colony or British Bechuanaland or Boer Republics or Zululand or Transvaal or Natalia Republic or Orange Free State).ti,ab. |
| 113 | or/11-112 [ALL SUB-SAHARAN AFRICA COUNTRIES] |
| 114 | 5 and 10 and 113 |

### S3. Table. CINAHL

| | |
|---|---|
| S15 | S7 AND S12 AND S13 AND S14 |
| S14 | Niger/ or (Niger not (Aspergillus or Peptococcus or Schizothorax or Cruciferae or Gobius or Lasius or Agelastes or Melanosuchus or radish or Parastromateus or Orius or Apergillus or Parastromateus or Stomoxys)).ti,ab. or Rwanda/ or (Rwanda or Ruanda).ti,ab. or Sierra Leone/ or (Sierra Leone or Salone).ti,ab. or Somalia/ or (Somalia or Somaliland).ti,ab. or south sudan/ or South Sudan.ti,ab. orTanzania/ or (Tanzania or Tanganyika or Zanzibar).ti,ab. or Togo/ or (Togo or Togolese Republic or Togoland).ti,ab. or Uganda/ or Uganda.ti,ab. or Angola/ or Angola.ti,ab. or Cameroon/ or (Cameroon or Kamerun or Cameroun).ti,ab. or Cape Verde/ or (Cape Verde or Cabo Verde).ti,ab. or Comoros/ or (Comoros or Glorioso Islands or Mayotte).ti,ab. or Congo/ or (Congo not ((Democratic Republic adj3 Congo) or congo red or crimean-congo)).ti,ab. or Cote d'Ivoire/ or (Cote d'Ivoire or Cote dIvoire or Ivory Coast).ti,ab. or eswatini/ or (eSwatini or Swaziland).ti,ab. or Ghana/ or (Ghana or Gold Coast).ti,ab. or Kenya/ or (Kenya or East Africa Protectorate).ti,ab. or Lesotho/ or (Lesotho or Basutoland).ti,ab. or Mauritania/ or Mauritania.ti,ab. or Nigeria/ or Nigeria.ti,ab. or "sao tome and principe"/ or (Sao Tome adj2 Principe).ti,ab. or Senegal/ or Senegal.ti,ab. or Sudan/ or (Sudan not South Sudan).ti,ab. or Zambia/ or (Zambia or Northern Rhodesia).ti,ab. or Zimbabwe/ or (Zimbabwe or Southern Rhodesia).ti,ab. or Botswana/ or (Botswana or Bechuanaland or Kalahari).ti,ab. or Equatorial Guinea/ or (Equatorial Guinea or Spanish Guinea).ti,ab. or Gabon/ or (Gabon or Gabonese Republic).ti,ab. or Mauritius/ or (Mauritius or Agalega Islands).ti,ab. or Namibia/ or (Namibia or German South West Africa).ti,ab. or South Africa/ or (South Africa or Cape Colony or British Bechuanaland or Boer Republics or Zululand or Transvaal or Natalia Republic or Orange Free State).ti,ab. |
| S13 | "Africa south of the Sahara"/ or ("Africa South of the Sahara" or sub-Saharan Africa or subSaharan Africa).ti,ab. or Central Africa.ti,ab. or Eastern Africa.ti,ab. or Southern Africa.ti,ab. or Western Africa.ti,ab. or Seychelles/ or Seychelles.ti,ab. or Benin/ or (Benin or Dahomey).ti,ab. or Burkina Faso/ or (Burkina Faso or Burkina Fasso or Upper Volta).ti,ab. or Burundi/ or (Burundi or Ruanda-Urundi).ti,ab. or Central African Republic/ or (Central African Republic or Ubangi-Shari).ti,ab. or Chad/ or Chad.ti,ab. or |

| | |
|---|---|
| | Democratic Republic Congo/ or (((Democratic Republic or DR) adj2 Congo) or Congo-Kinshasa or Belgian Congo or Zaire or Congo Free State).ti,ab. or Eritrea/ or Eritrea.ti,ab. or Ethiopia/ or (Ethiopia or Abyssinia).ti,ab. or Gambia/ or Gambia.ti,ab. or Guinea/ or (Guinea not (New Guinea or Guinea Pig* or Guinea Fowl or Guinea-Bissau or Portuguese Guinea or Equatorial Guinea)).ti,ab. or Guinea-Bissau/ or (Guinea-Bissau or Portuguese Guinea).ti,ab. or Liberia/ or Liberia.ti,ab. or Madagascar/ or (Madagascar or Malagasy Republic).ti,ab. or Malawi/ or (Malawi or Nyasaland).ti,ab. or Mali/ or Mali.ti,ab. or Mozambique/ or (Mozambique or Mocambique or Portuguese East Africa).ti,ab. |
| S12 | S8 OR S9 OR S10 OR S11 |
| S11 | mental health or (mental adj2 wellness) or mental disorder* or psychological health or emotional health or mental fitness or psychosocial well-being or behavio* health or psychiatr* wellness or Lunac* or melancholia or hyster* or nervous breakdown or madness or moral insanity or bipolar disorder* or psychotic disorder or anxiety or anxiety disorder* or depression or schizophreni* |
| S10 | psychiatry/ |
| S9 | mental disorder/ |
| S8 | mental health/ |
| S7 | S1 OR S2 OR S3 OR S4 OR S5 OR S6 |
| S6 | (pathway* to care or clinical pathway*) or (health adj2 behavio?r) or access to care or patient journey or healthcare pathway* or service access or care continuum or care routes or treatment at |
| S5 | ((pathway* to care or clinical pathway* or (health adj2 behavio?r) or access to care or patient journey or healthcare pathway* or service access or care continuum or care routes or treatment) adj2 access) |
| S4 | access to care or access to healthcare or access to services |
| S3 | access to care/ |
| S2 | health seeking behavior/ |
| S1 | clinical pathway/ or pathway* to care or patient care/ |

S4. Table. Global Index Medicus – world Health Organization

| | |
|---|---|
| | clinical pathway/ or pathway* to care or patient care/ or health seeking behavior/ or access to care/ or access to care or access to healthcare or access to services or ((pathway* to care or clinical pathway* or (health adj2 behavio?r) or access to care or patient journey or healthcare pathway* or service access or care continuum or care routes or treatment) adj2 access) |
| | mental health/ or mental disorder/ or psychiatry/ or mental health or (mental adj2 wellness) or mental disorder* or psychological health or emotional health or mental fitness or psychosocial well-being or behavio* health or psychiatr* wellness or Lunac* or melancholia or hyster* or nervous breakdown or madness or moral insanity or bipolar disorder* or psychotic disorder or anxiety or anxiety disorder* or depression or schizophreni* |
| AND | "Africa south of the Sahara"/ or ("Africa South of the Sahara" or sub-Saharan Africa or subSaharan Africa).ti,ab. or Central Africa.ti,ab. or Eastern Africa.ti,ab. or Southern Africa.ti,ab. or Western Africa.ti,ab. or Seychelles/ or Seychelles.ti,ab. or Benin/ or (Benin or Dahomey).ti,ab. or Burkina Faso/ or (Burkina Faso or Burkina Fasso or Upper Volta).ti,ab. or Burundi/ or (Burundi or Ruanda-Urundi).ti,ab. or Central African Republic/ or (Central African Republic or Ubangi-Shari).ti,ab. or Chad/ or Chad.ti,ab. or Democratic Republic Congo/ or (((Democratic Republic or DR) adj2 Congo) or Congo-Kinshasa or Belgian Congo or Zaire or Congo Free State).ti,ab. or Eritrea/ or Eritrea.ti,ab. or Ethiopia/ or (Ethiopia or Abyssinia).ti,ab. or Gambia/ or Gambia.ti,ab. or Guinea/ or (Guinea not (New Guinea or Guinea Pig* or Guinea Fowl or Guinea-Bissau or Portuguese Guinea or Equatorial Guinea)).ti,ab. or Guinea-Bissau/ or (Guinea-Bissau or Portuguese Guinea).ti,ab. or Liberia/ or Liberia.ti,ab. or Madagascar/ or (Madagascar or Malagasy Republic).ti,ab. or Malawi/ or (Malawi or Nyasaland).ti,ab. or Mali/ or Mali.ti,ab. or Mozambique/ or (Mozambique or Mocambique or Portuguese East Africa).ti,ab. or Niger/ or (Niger not (Aspergillus or Peptococcus or Schizothorax or Cruciferae or Gobius or Lasius or Agelastes or Melanosuchus or radish or |

| | |
|---|---|
| | Parastromateus or Orius or Apergillus or Parastromateus or Stomoxys)).ti,ab. or Rwanda/ or (Rwanda or Ruanda).ti,ab. or Sierra Leone/ or (Sierra Leone or Salone).ti,ab. or Somalia/ or (Somalia or Somaliland).ti,ab. or south sudan/ or South Sudan.ti,ab. orTanzania/ or (Tanzania or Tanganyika or Zanzibar).ti,ab. or Togo/ or (Togo or Togolese Republic or Togoland).ti,ab. or Uganda/ or Uganda.ti,ab. or Angola/ or Angola.ti,ab. or Cameroon/ or (Cameroon or Kamerun or Cameroun).ti,ab. or Cape Verde/ or (Cape Verde or Cabo Verde).ti,ab. or Comoros/ or (Comoros or Glorioso Islands or Mayotte).ti,ab. or Congo/ or (Congo not ((Democratic Republic adj3 Congo) or congo red or crimean-congo)).ti,ab. or Cote d'Ivoire/ or (Cote d'Ivoire or Cote dIvoire or Ivory Coast).ti,ab. or eswatini/ or (eSwatini or Swaziland).ti,ab. or Ghana/ or (Ghana or Gold Coast).ti,ab. or Kenya/ or (Kenya or East Africa Protectorate).ti,ab. or Lesotho/ or (Lesotho or Basutoland).ti,ab. or Mauritania/ or Mauritania.ti,ab. or Nigeria/ or Nigeria.ti,ab. or "sao tome and principe"/ or (Sao Tome adj2 Principe).ti,ab. or Senegal/ or Senegal.ti,ab. or Sudan/ or (Sudan not South Sudan).ti,ab. or Zambia/ or (Zambia or Northern Rhodesia).ti,ab. or Zimbabwe/ or (Zimbabwe or Southern Rhodesia).ti,ab. or Botswana/ or (Botswana or Bechuanaland or Kalahari).ti,ab. or Equatorial Guinea/ or (Equatorial Guinea or Spanish Guinea).ti,ab. or Gabon/ or (Gabon or Gabonese Republic).ti,ab. or Mauritius/ or (Mauritius or Agalega Islands).ti,ab. or Namibia/ or (Namibia or German South West Africa).ti,ab. or South Africa/ or (South Africa or Cape Colony or British Bechuanaland or Boer Republics or Zululand or Transvaal or Natalia Republic or Orange Free State).ti,ab. |

### S5. Table. Selection criteria

| Category | Inclusion | Exclusion |
|---|---|---|
| Article type | Peer-reviewed articles, research studies, empirical studies, grey literature (dissertations and theses). | Narrative/scoping reviews, opinion/correspondence/commentary articles, pre-prints, abstracts, and conference presentations. |
| Study type and design | Qualitative studies (case study analysis, discourse analysis, focus group discussion-based, interview-based, action research, grounded theory, observation, participant observation); mixed-method studies where qualitative evidence is epistemologically distinct from and reported separately to quantitative evidence. | Quantitative studies, qualitative process evaluations, realist evaluations, study protocols |
| Environment/setting | SSA in the context of the local mental health system, regulatory, and policymaking settings in the same environments, research, and development. | North Africa, LMICs that are not part of SSA, High-income countries in the context of the local mental health system, regulatory and policymaking settings that pertain to mental health services in high-income countries, primarily. |
| Perspective | Stakeholder perspective, e.g., mental health researchers and healthcare workers, regulators, policymakers, health managers, formal health service providers, informal health service providers, partner organizations, and caregivers. | Studies in which mental health service delivery is not the primary focus. |
